# Acute effect of high-intensity interval training on fetal blood flow distribution

**DOI:** 10.64898/2026.05.27.26354197

**Authors:** HMS Skarstad, S Skrede, KL Haganes, ER Ashby, MAJ Sujan, K Deibele, H Mørch, G Haugen, KÅ Salvesen, T Moholdt

## Abstract

**Objectives:** To examine the acute effects of a single bout of high-intensity interval training (HIIT) on fetal blood flow distribution during the third trimester of pregnancy.

**Methods:** Thirty-four healthy pregnant participants (mean age 31.6 years, standard deviation (SD) 4.1; gestational week 33.8 (SD 0.4) completed eight 30-second high-intensity cycling work-bouts interspersed with 2-minute rest periods. Fetal heart rate (FHR), maternal blood pressure, and Doppler-derived blood flow indices in the middle cerebral artery, umbilical artery and vein, and ductus venosus were assessed before and after exercise. We estimated fetal liver blood flow and the ratio of umbilical vein flow to ductus venosus. Maternal heart rate (HR) and FHR were recorded throughout exercise. Paired *t*-tests compared pre- and post-exercise values.

**Results:** No significant changes were observed in fetal blood flow indices or distribution following exercise. Average maternal HR and FHR during the work-bouts were 158 bpm (SD 16) and 152 bpm (SD 12), respectively. Following HIIT, maternal systolic blood pressure increased by 5 mmHg (95% CI 1 to 8, *p*=.014), maternal HR by 22 bpm (95% CI 15 to 28, *p*<.001), and FHR by 13 bpm (95% CI 10 to 17, *p*<.001). We recorded 16 instances of FHR above normal range during HIIT.

**Conclusion:** A single HIIT session in late pregnancy increased maternal blood pressure and HR and transiently elevated FHR but did not affect fetal blood flow indices or distribution. Brief episodes of fetal tachycardia were observed but appeared to be clinically insignificant. Future research should investigate the effects of repeated HIIT exposure during pregnancy.

**What is already known on this topic:**

- Moderate-intensity exercise during pregnancy is considered safe and beneficial for maternal and fetal health.
- High-intensity exercise during pregnancy may transiently affect fetal heart rhythm, including brief episodes of fetal tachycardia or arrhythmia.

**What this study adds:**

- This study provides a comprehensive assessment of the acute effects of a single high-intensity interval training (HIIT) session on fetal blood flow indices and blood flow distribution, including fetal liver perfusion, during pregnancy.
- A single HIIT session did not alter fetal blood flow measurements despite transient increases in fetal heart rate.

**How this study might affect research, practice or policy:**

- Evidence regarding the safety of HIIT during pregnancy remains limited.

These findings suggest that acute HIIT does not adversely affect fetal blood flow parameters in healthy pregnancies and may support inclusion of HIIT as a time-efficient exercise option during pregnancy

## Introduction

Current guidelines recommend that pregnant individuals engage in at least 150 minutes of moderate-intensity physical activity weekly (1, 2). Regular exercise is associated with reduced incidence of pregnancy complications such as gestational diabetes and pre-eclampsia and improves maternal mental well-being (2-4). Despite these benefits, few pregnant individuals meet current physical activity recommendations (5). Given the global rise in prevalence of overweight and obesity, alongside declining physical activity levels among reproductive-age females, promoting exercise during pregnancy is increasingly important (3, 6, 7).

A common barrier to exercise is lack of time (4). High-intensity interval training (HIIT), which consists of brief bouts of high-intensity exercise interspersed with recovery periods, may offer a time-efficient alternative to traditional exercise modalities (8). HIIT has demonstrated beneficial cardiometabolic effects in women of reproductive age with overweight and obesity (9), and both pregnant and non-pregnant individuals report high levels of enjoyment during HIIT participation (10, 11). Its efficiency and perceived enjoyment may therefore support greater adherence to physical activity during pregnancy.

Despite these potential advantages, concerns regarding fetal safety remain a major barrier to exercise participation during pregnancy (4). While moderate-intensity exercise is widely considered safe for both mother and fetus (7, 12, 13), there is limited evidence regarding high-intensity exercise. Previous studies investigating HIIT in pregnancy have reported mixed findings, with some demonstrating good maternal and fetal tolerance (10, 14-16), others have suggested transient alterations in fetoplacental circulation (17, 18).

The HITFLOW study aimed to address this knowledge gap by evaluating acute changes in fetal blood flow distribution following a single HIIT session during the third trimester of pregnancy. We hypothesized that a single session of HIIT would not alter fetal blood flow and would be well tolerated by the fetus.

## Methods

### Trial design

HITFLOW was an experimental laboratory study investigating fetal blood flow before and after one HIIT session during the third trimester of pregnancy. The study was conducted at the Norwegian University of Science and Technology (NTNU) and the Department of Obstetrics and Gynaecology at St. Olav’s University Hospital, Trondheim, Norway. The trial was registered in Clinical Trials (clinicaltrials.gov, ID: NCT04288479) and approved by the Norwegian Regional Committees for Medical and Health Research Ethics of South-Eastern Norway (reference 62993). Recruitment commenced in April 2022 following delays related to the COVID-19 pandemic, and the final participant was enrolled in April 2025.

### Participants

We recruited pregnant individuals aged ≥18 years with singleton pregnancies between gestational week 32 and 36. Individuals of all fitness levels were eligible to participate. Exclusion criteria included pre-existing or gestational hypertension, gestational diabetes mellitus, and any condition preventing participation in cycling exercise. Participants were recruited through advertisements on social media, university web pages, and posters at St. Olav’s University Hospital. Eligibility was assessed by telephone before inclusion. All participants provided written informed consent prior to participation.

### HIIT training protocol

Participants attended the outpatient clinic at the Department of Obstetrics and Gynaecology following a ≥2 hours fast. We performed ultrasound assessments and measurements of maternal blood pressure (BP) and heart rate (HR) before and after the training session. Exercise was undertaken on a stationary bike (Impulse Spinning G2, Impulse Fitness,). Following a 10-minute warm-up, the participants completed eight 30-second high-intensity work-bouts separated by 2-minute recovery periods (Figure 1). The target intensity was 80-90% of estimated maximal HR (HR_max_), calculated using the age-based prediction formula: 211–0.64×age (19). Perceived exertion was recorded after each work-bout using the Borg 6-20 scale (20). Maternal HR was continuously monitored using a chest strap (Polar H10, Polar Electro, Finland) connected to the Polar Beat application. Participants could either rest completely or pedal slowly during recovery periods and were free to sit or stand during work-bouts. We assessed fetal heart rate (FHR) immediately after each work-bout using a handheld Doppler device (SonicaidOne, Huntleigh Healthcare, United Kingdom).

**Figure 1.**
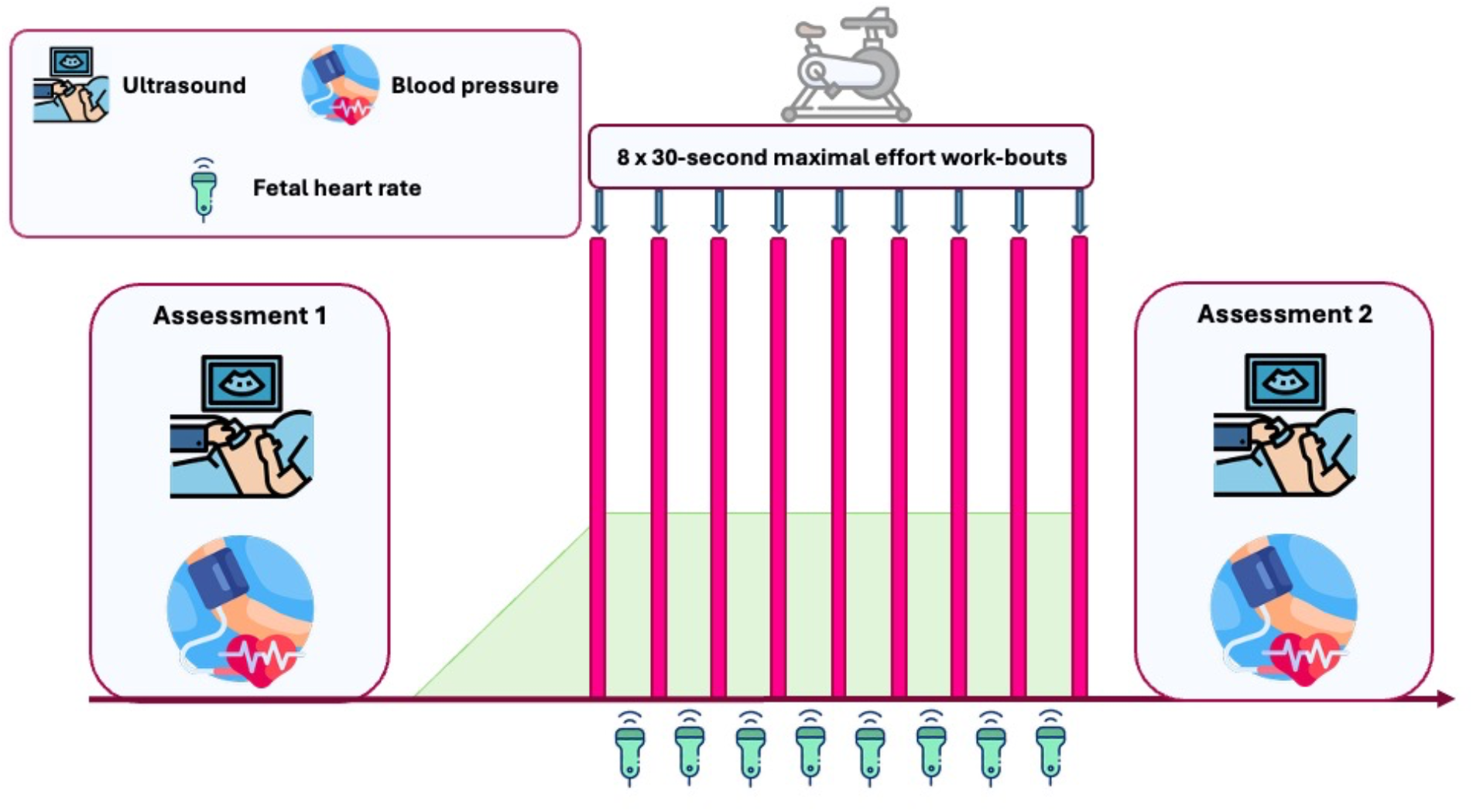
Study design. Participants underwent ultrasound and blood pressure assessments before and after a single high-intensity interval training (HIIT) session performed on an exercise bicycle. Following a 10-minute warm-up, participants completed eight 30-second high-intensity work-bouts separated by 2-minute recovery periods. We performed the post-exercise ultrasound and blood pressure assessment immediately after the final work-bout. After the assessments, participants underwent body composition analysis and completed a questionnaire regarding physical activity, medication use, and parity.

### Outcomes

#### Primary outcome

The primary outcome was changes in fetal blood flow following a single HIIT session. We assessed blood flow using Doppler ultrasound immediately before and after exercise using a Voluson E10 and later Voluson Expert 22 ultrasound system, equipped with C2-9 and RM7C probes (GE HealthCare, United States). Participants were in a supine or slightly reclined position during examinations and fasted ≥2 hours prior to assessments. An experienced obstetrician trained in fetal medicine performed the ultrasound assessments. We followed the International Society of Ultrasound in Obstetrics and Gynaecology practice guidelines for use of Doppler velocimetry in obstetrics (21). Ultrasound procedures were performed according to the “as low as reasonably achievable”-principle for ultrasound exposure (22). The insonation angle was kept as close to 0º as possible and never exceeded 30°. At least three consecutive measurements were obtained for all Doppler variables.

We assessed blood flow variables of the umbilical artery (UA), umbilical vein (UV), middle cerebral artery (MCA), and ductus venosus (DV). UA measurements were obtained from a free cord loop in the mid-portion of the umbilical cord (21, 23). MCA measurements were obtained in an axial section of the fetal brain including the thalami and the sphenoid wings with the pulsed Doppler gate positioned in the proximal third of the MCA. We avoided excess pressure on the fetal head during examination. Peak systolic velocity was set to be the highest point of the waveform. Pulsatility index (PI) in the MCA was calculated using peak-to-peak height of waveforms and mean height over one cardiac cycle (24). PI in the umbilical cord vessels was calculated automatically during measurements (23).

Internal vessel diameters of the UV (D_UV_) and DV (D_DV_) were measured from the inner vessel walls, perpendicular to the vessel axis and averaged across three diameter measurements (25). Time-averaged maximum velocity (TAMXV) was measured in a longitudinal section in a straight portion of the intra-abdominal UV and at the inlet of the DV (26) and calculated as the average of at least three repeated measurements. We calculated blood flow (Q) in the UV and DV as: *Q* = *h* × (*D*/2)^2^ × *π* × *TAMXV* where *h* represents the coefficient for spatial blood velocity profile (0.5 for UV 0.5 and 0.7 for DV), and D is vessel diameter (27). We calculated fetal liver blood flow (Q_liver_) as UV flow minus DV flow (Q_UV_ - Q_DV_), and the proportion of UV flow shunted through DV (DV_ratio_) as Q_DV_/Q_UV_.

We measured FHR before and after exercise using Doppler tracings in the UA and during the training session using a handheld Doppler device (SonicaidOne, Huntleigh Healthcare, United Kingdom). Exercise was discontinued if FHR decreased below 110 bpm or if FHR ≥180 bpm persisted beyond the recovery period, in which case participants were referred for same-day fetal assessment at the Centre for Fetal Medicine at St. Olav’s Hospital. Fetal biometric measurements, including biparietal diameter, middle abdominal diameter, and femur length, were recorded before exercise.

#### Secondary outcomes

We measured maternal BP and resting HR in a supine position immediately before and after the HIIT session using an automated BP machine (Welch Allyn, Germany). Three measurements were obtained at 1-minute intervals, and we report the mean values. We recorded maternal HR at the end of each work-bout using the Polar Beat application. We estimated body composition, including body weight, body mass index (BMI), muscle mass, and fat mass, after the final ultrasound examination using bioelectrical impedance analysis (Inbody 720, Biospace CO, Korea). Participants were measured while fasting, barefoot, and wearing light clothing. Participants completed a questionnaire regarding physical activity habits, medication use, parity, and estimated due date. Delivery outcomes, including birth weight, birth length, head circumference, placenta weight, mode of delivery, and adverse events, were obtained from electronic hospital records.

#### Sample size

Owing to the exploratory nature of the study, no formal sample-size calculation was performed. We aimed to recruit minimum 30 participants to provide preliminary data on fetal blood flow variability. Ultimately, we enrolled 34 participants, consistent with recommended sample sizes for pilot and feasibility studies (24-50 participants) (28).

#### Statistical methods

We compared pre- and post-exercise values using paired *t*-tests. Variables with non-normal distributions were log-transformed prior to analysis, and results are presented as percentage change with corresponding confidence intervals (CI) and *p*-values. For variables that remained non-normally distributed after transformation, we used Wilcoxon signed-rank test and report median values (IQR), *p*-values, and *r*-values. We adjusted PI according to FHR using repeated analysis of covariance. Continuous variables recorded at a single point in time are presented as means with standard deviations (SD). We defined statistical significance as *p*<0.05. Owing to the exploratory nature of the study, we did not adjust for multiple comparisons. Statistical analyses were conducted in IBM SPSS Statistics, version 30.0.

#### Patient and public involvement

We did not involve patients or the public in the design, conduct, reporting, or dissemination plans for this research.

## Results

We included 34 participants. Although we initially aimed to recruit 30 participants, we enrolled four additional participants because of high interest in participation. We tested the first participant on 4 April 2022 and the final participant on 23 April 2025. Participants had a mean age of 31.6 years (SD 4.1), mean BMI of 27.8 kg/m^2^ (SD 3.4), and the mean gestational age of 33.8 weeks (SD 1.5). All fetal biometric measurements fell within the normal ranges. Most participants reported exercising during the past two weeks (Table 1). Mean infant birth weight was 3481 g (SD 403), and mean birth length was 49.8 cm (SD 1.9) (Supplementary table 3). Twenty-four participants (82%) delivered vaginally, five (15%) by caesarean section, and one (3%) by vaginal breech delivery. One participant delivered preterm at gestational length 36+1 weeks. Six other adverse birth events were recorded, including placenta previa, low Apgar score, retained placenta and prolonged labour.

**Table 1.**
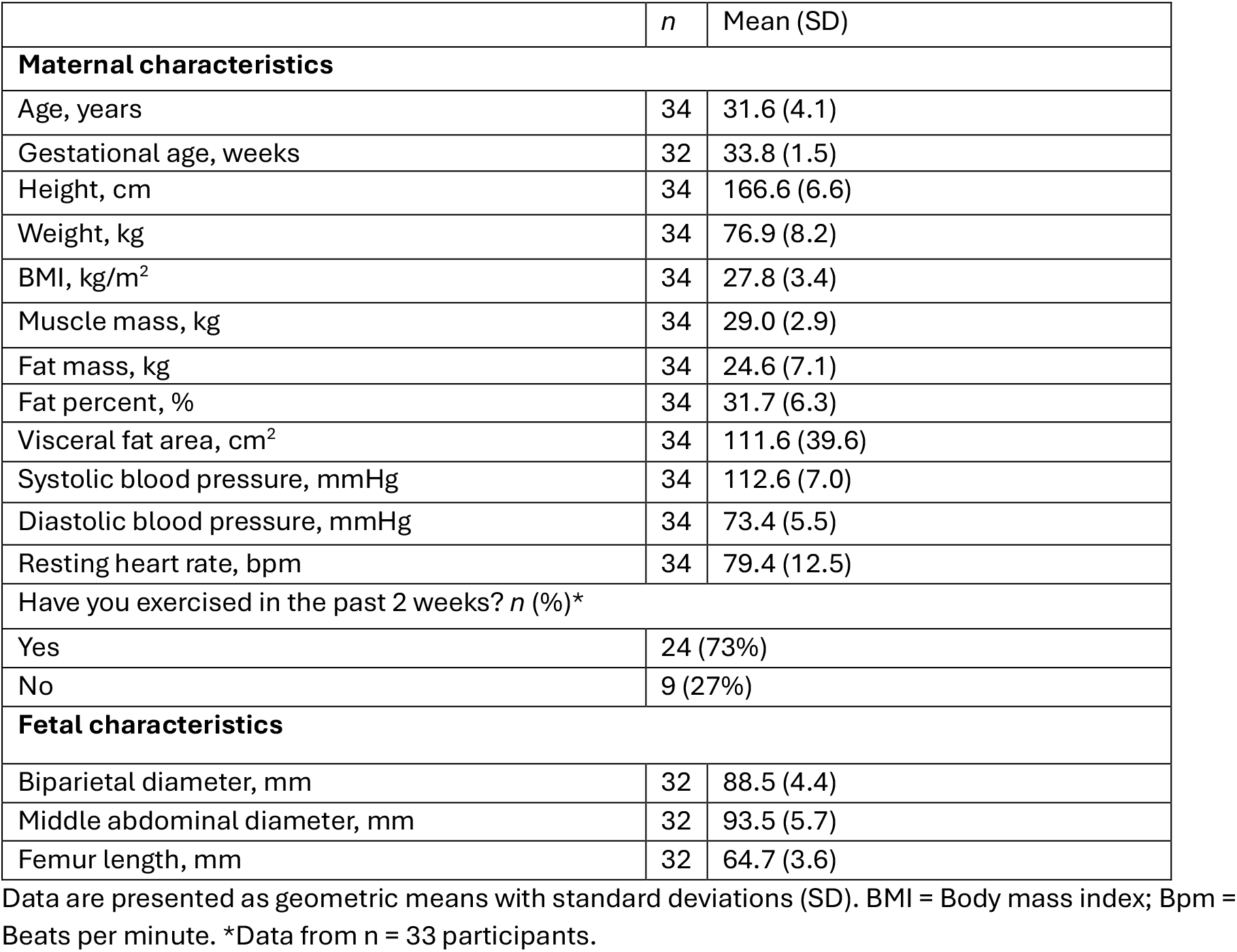
Participant characteristics.

Incomplete ultrasound recordings prevented calculation of fetal liver blood flow and DV_ratio_, and some fetal biometry measurements in a small number of participants. Technical issues with HR monitoring equipment also caused some missing maternal and fetal HR data during exercise.

### Maternal and fetal response to HIIT

Table 2 shows maternal and fetal blood flow parameters before and after exercise. HIIT did not change blood flow, PI, vessel diameter, or flow velocity in any of the examined vessels (Figure 2). Similarly, HIIT did not affect estimated fetal liver blood flow or DV_ratio_. Although UA PI showed a non-significant tendency of reduction after exercise, PI values remained unchanged after adjustment for FHR. FHR increased significantly from 141 bpm (SD 8) before exercise to 155 bpm (SD 10) after exercise. Mean FHR during exercise was 152 bpm (SD 12). Five participants experienced transient fetal tachycardia (FHR ≥180 bpm), including one with a peak FHR ≥200 bpm (Figure 3). All episodes resolved spontaneously without intervention. Mean FHR variability during exercise was 26 bpm (SD 13) (Supplementary Table 2). Figure 4 shows maternal BP and resting HR before and after exercise. HIIT increased systolic BP and resting HR, whereas diastolic BP remained unchanged (Table 2). During the session, participants reached a mean exercise intensity corresponding to 84% (SD 5) of estimated HR_max_. Supplementary Table 2 presents complete exercise data.

**Table 2.**
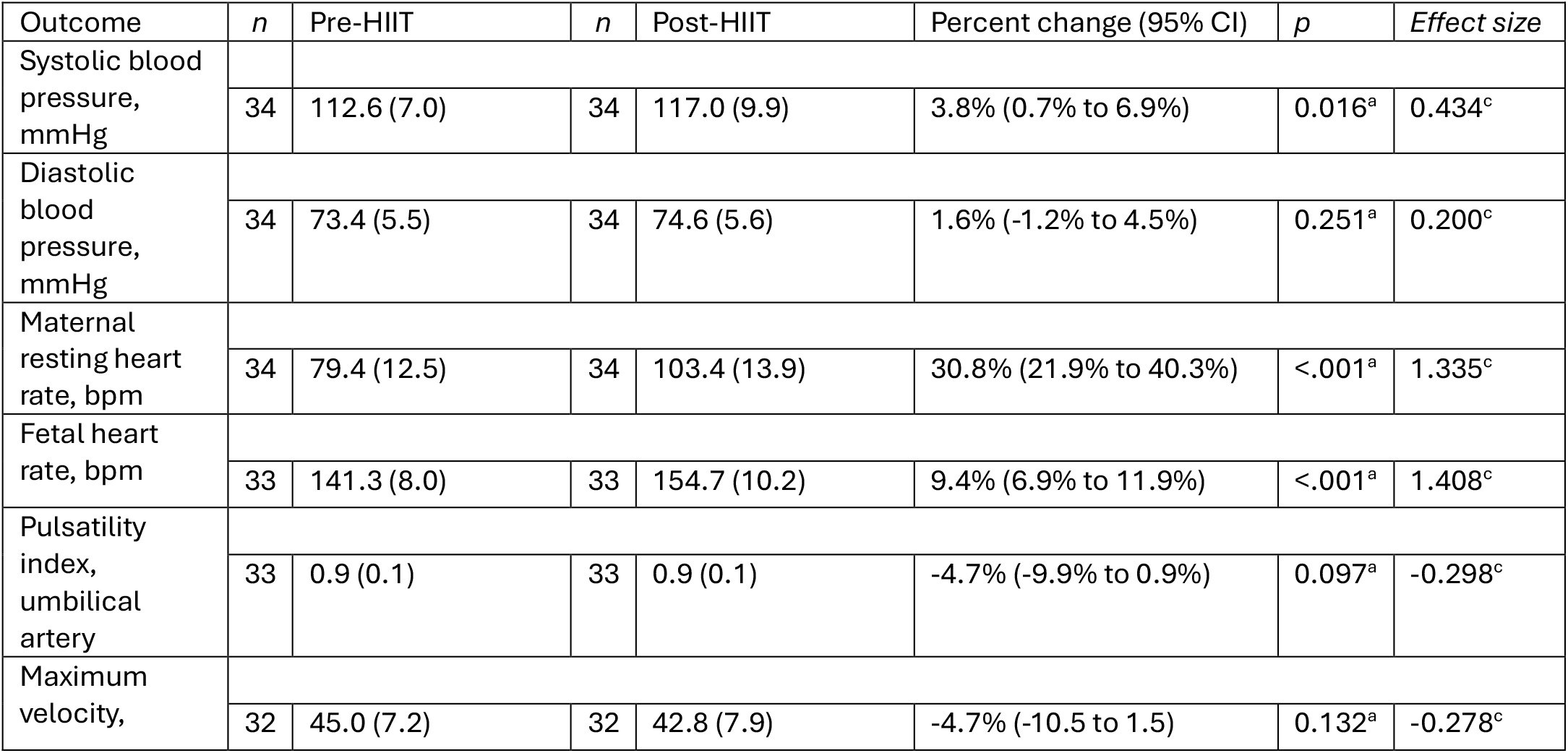
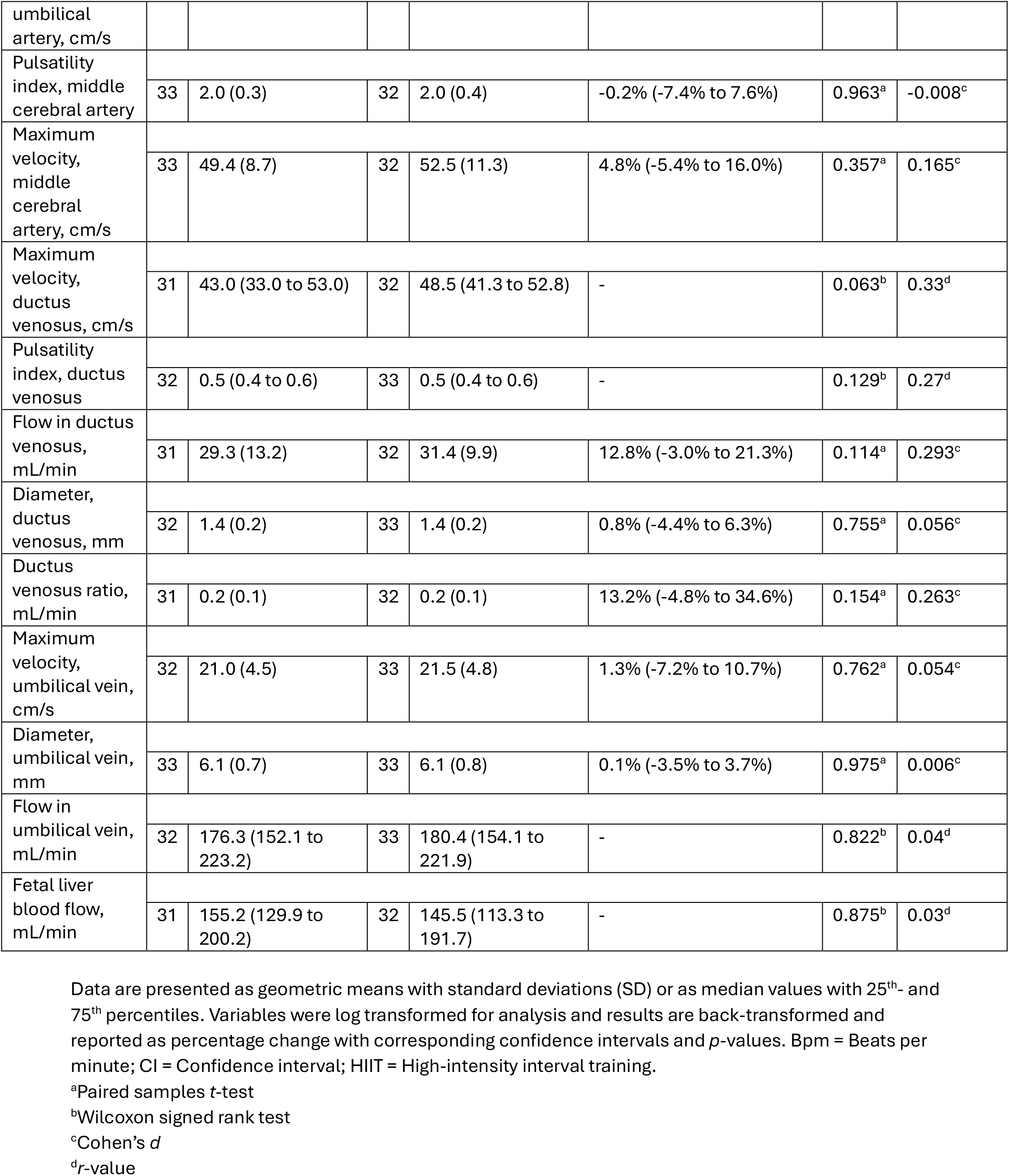
Maternal-fetal blood flow parameters before and after exercise.

**Figure 2.**
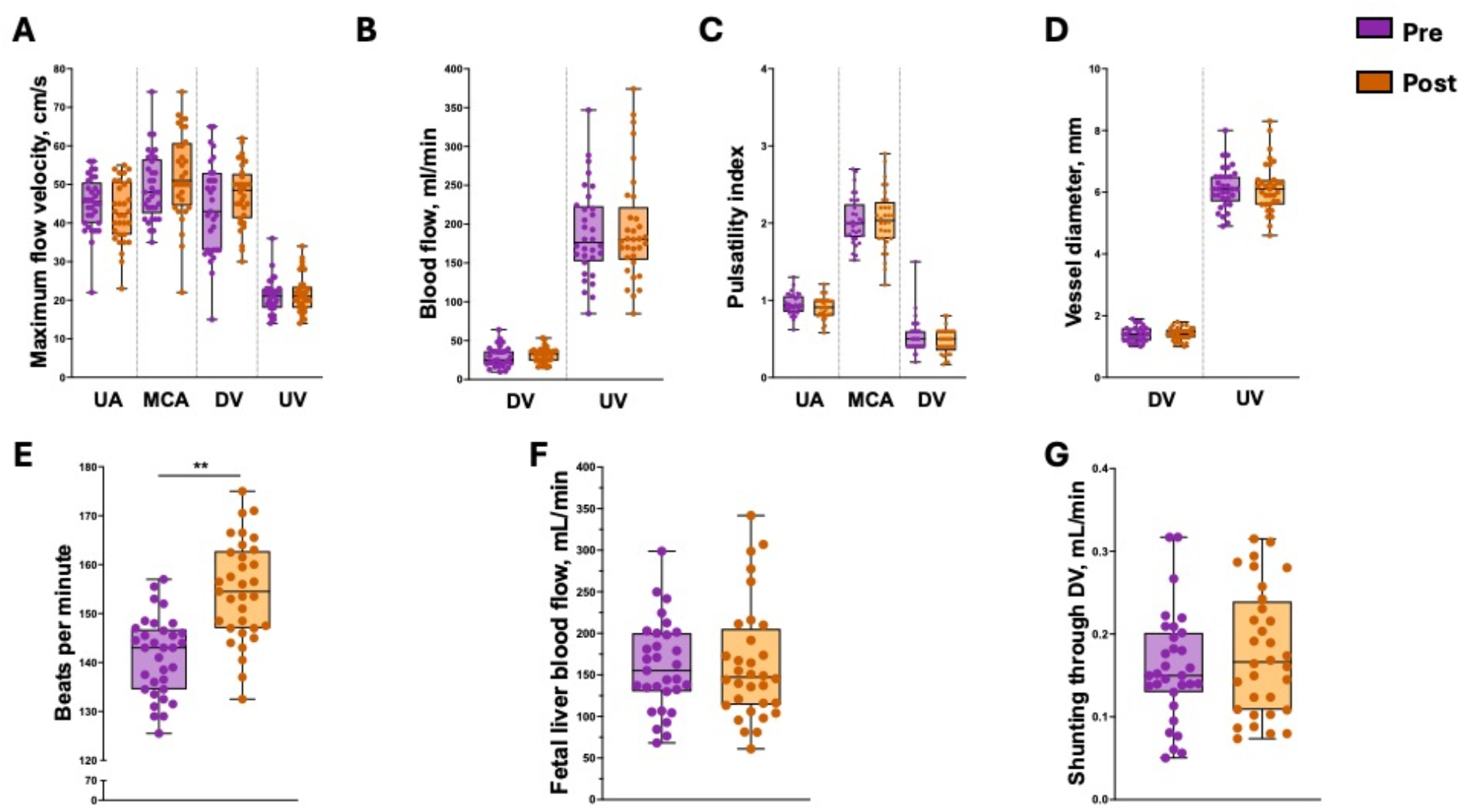
Blood flow parameters. Doppler ultrasound measurements obtained before (pre) and after (post) exercise. **A)** Maximum flow velocity in the umbilical artery (UA), middle cerebral artery (MCA), ductus venosus (DV), and umbilical vein (UV). **B)** Blood flow through the DV and UV. **C)** Pulsatility index (PI) in the UA, MCA, and DV. **D)** Vessel diameter in the DV and UV. **E)** Fetal heart rate in beats per minute (bpm). **F)** Estimated fetal liver blood flow. **G)** Umbilical venous blood flow shunted through the DV. Data are presented as individual data points with mean and SD. *P*-value were calculated using paired *t*-tests or Wilcoxon signed rank test. \*\**P*<.001.

**Figure 3.**
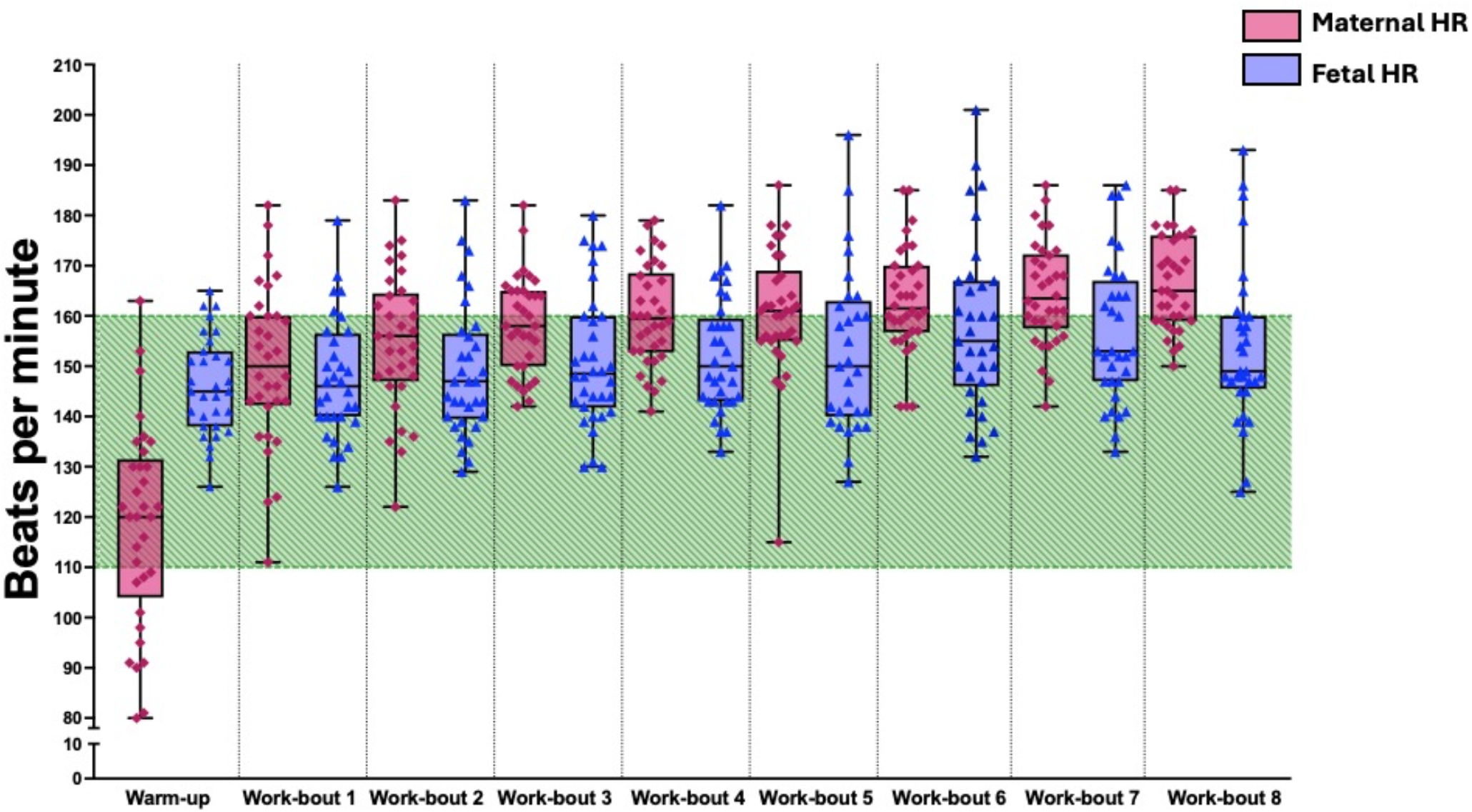
Maternal and fetal heart rate during exercise. Maternal heart rate (HR) was recorded continuously during each work-bout, and fetal heart rate (FHR) was measured immediately at the start of the recovery period. The shaded area indicates normal fetal heart rate range. Five participants experienced transient fetal tachycardia during exercise, including one participant with a peak HR ≥ 200 beats per minute (bpm). Data are presented as individual data points with mean and SD for each work-bout.

**Figure 4.**
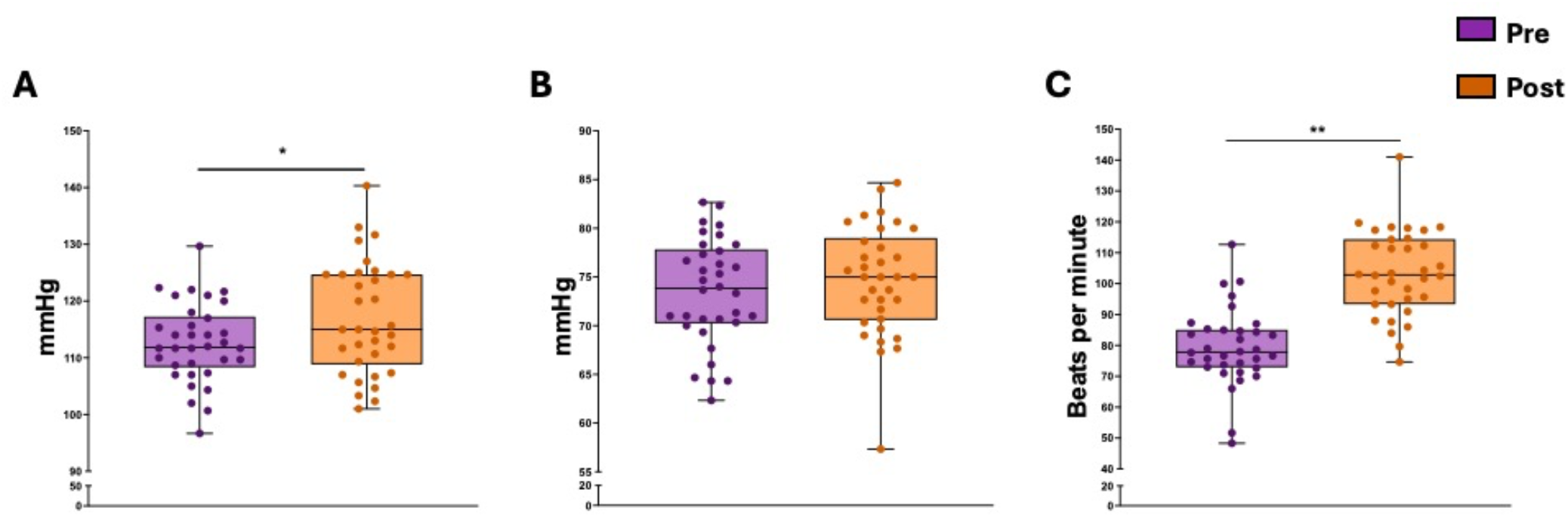
Maternal cardiovascular responses before and after exercise. **A)** Systolic blood pressure. **B)** Diastolic blood pressure. **C)** Resting heart rate. Data are presented as individual data points with mean and SD before and after exercise. *P*-value were calculated using paired t-tests. \**P*<.05, \*\**P*<.001.

### Adverse events

Two participants experienced mild dizziness and nausea during the ultrasound examinations, one during the pre-exercise assessment and one during the post-exercise assessment. Both episodes resolved spontaneously after a brief pause. We recorded no other adverse events during the study.

## Discussion

This study showed that a single session of HIIT during the third trimester did not alter fetal blood flow distribution. Although FHR increased during exercise and five participants experienced brief episodes of fetal tachycardia, all Doppler assessments before and after exercise remained within normal ranges, and we observed no signs of fetal distress. Maternal systolic BP and HR increased after exercise, consistent with expected physiological responses to acute high-intensity exercise.

Transient FHR elevations ≥180 bpm occurred in five participants but resolved spontaneously without intervention. Although FHR increased after exercise, mean FHR remained within the normal reference range (110-160 bpm), consistent with previous studies (13, 17, 29). FHR often parallels maternal HR during exercise (13, 30), and mild tachycardia may represent a normal fetal adaptation to maternal exertion (31). Importantly, fetal tachycardia is rarely clinically significant unless FHR exceeds 200 bpm or persists for >2 minutes (13, 32). Exercise protocol characteristics may influence fetal responses to HIIT. Participants in our study completed eight 30-second work-bouts separated by 2-minute recovery periods, similar to protocols previously reported to be well tolerated during pregnancy (16, 33). In contrast, studies reporting fetal arrhythmias have typically used longer high-intensity work-bouts lasting ≥5-minute. Prolonged high-intensity exercise may therefore increase the likelihood of transient fetal rhythm disturbances.

Previous studies have also suggested that fetal arrhythmias occur more frequently when maternal HR exceeds 90% of HR_max_ during exercise (17, 18, 34). Exercise intensities between 80-90% of HR_max_ generally appear well tolerated and do not consistently affect FHR or fetal blood flow (16, 33), and even intensities above 90% of HR_max_ may be tolerated in healthy pregnancies (10). Although participants in our study were instructed to exercise at 80-90% of estimated HR_max_, some exceeded this range without adverse effects, while others experienced FHR ≥180 bpm at lower maternal HRs. These findings may reflect limitations of age-predicted HR_max_ formulas during pregnancy (35, 36).

Participant fitness level may also influence fetal responses to high-intensity exercise. Studies reporting fetal arrhythmias have often included elite athletes or highly trained individuals who may achieve higher HRs and sustain them for longer periods (17, 18, 34). In contrast, studies including participants with mixed fitness levels have generally reported good fetal tolerance to HIIT (10). Although we included participants regardless of fitness level, most reported regular exercising regularly, suggesting that our sample may represent a relatively active subgroup compared with the general pregnant population.

Unlike previous studies, we assessed blood flow in multiple fetal vessels, including the UA, MCA, UV, and DV, and estimated fetal liver blood flow and DV_ratio_. To our knowledge, this is the first study to examine the acute effect of HIIT on fetal blood flow distribution and fetal liver perfusion during pregnancy. Although increased blood flow to the fetal liver has been associated with greater neonatal fat mass (26), no previous studies have evaluated its response to maternal exercise.

Our findings align with previous literature suggesting that exercise has limited effects on fetoplacental circulation. The UA is the most frequently studied vessel during maternal exercise, although findings remain inconsistent. Some studies have reported reduced UA PI immediately after HIIT, suggesting increased fetoplacental perfusion (10, 33), whereas others observed no change or transient increases in PI during high-intensity exercise (17, 18). A systematic review concluded that maternal exercise generally does not affect UA blood flow indices (13). We similarly observed no significant changes in UA PI or other fetal blood flow measurements following HIIT.

We did not assess uterine artery blood flow and therefore cannot comment on uteroplacental perfusion during exercise. One previous study reported transient reductions in uterine artery volume flow of 60-80% during high-intensity exercise, with reductions persisting for up to 10 minutes after exercise cessation (18). We also only performed three repeated vessel diameter measurements, which may have reduced measurement precision (37).

This study primarily evaluated the acute safety and feasibility of HIIT during late pregnancy. Although we observed no adverse changes in fetal blood flow distribution following a single exercise session, long-term effects of repeated HIIT exposure during pregnancy remain unknown. Moderate-intensity exercise training during pregnancy promotes vascular remodelling, placental development, and endothelial function (38), but few studies have examined whether HIIT produces similar adaptations. To our knowledge, only two trials have investigated repeated HIIT during pregnancy, and neither included detailed assessments of fetal well-being (15, 39). To our knowledge, few previous trials have investigated repeated HIIT during pregnancy, and none have included detailed assessments of fetal well-being (15, 39).

The increases in maternal systolic BP and HR after exercise likely reflect normal physiological responses to acute exertion. We obtained baseline measures after approximately 30 minutes of rest, whereas we performed post-exercise assessments occurred immediately after the final work-bout. Acute exercise increases cardiac through increased stroke volume and HR output to meet metabolic demands (40), and pregnancy-related cardiovascular adaptations may further augment these responses (36).

### Limitations

This study has several limitations. First, we did not assess uterine artery blood flow, which limited our ability to evaluate uteroplacental perfusion during exercise. Second, we cannot determine long-term effects of repeated HIIT exposure during pregnancy because we only evaluated acute responses to a single exercise session. Third, we obtained fewer repeated vessel diameter measurements than commonly recommended, which may have reduced precision. Technical challenges with HR monitoring also resulted in some missing data and potential measurement artefacts. Finally, our relatively small sample consisted primarily of healthy and physically active participants which may limit generalisability to less active populations, multifetal pregnancies, and pregnancies complicated with comorbidities.

### Conclusions and clinical implications

A single HIIT session during late pregnancy did not alter fetal blood flow distribution. Although some participants experience transient fetal tachycardia during exercise, all episodes resolved spontaneously, and we observed no evidence of fetal compromise. These findings support the short-term tolerability of HIIT in healthy pregnancies.

Most pregnant individuals do not achieve recommended physical activity levels during pregnancy, highlighting the need for effective and time-efficient exercise strategies. HIIT may represent one such approach, although additional research is needed before firm recommendations can be made. Future studies should investigate the effects of repeated HIIT exposure during pregnancy and compare different exercise intensities, work-bout durations, and training frequencies to identify safe and effective HIIT protocols for pregnant populations.

## Data Availability

The datasets used during the current study are currently not publicly available but can be obtained from the principal investigator (Trine Moholdt) upon reasonable request.

## Declarations

### Ethics approval and consent to participate

The HITFLOW trial was registered in Clinical Trials (clinicaltrials.gov, ID: NCT04288479) and approved by Norwegian Regional Committees for Medical and Health Research Ethics (ID: 62993). The research was performed according to the Helsinki Declaration. All participants provided written consent prior to participating in the trial. We gave information on voluntary participation and that the participants were free to withdraw their consent at any time. We collected the signed consent form from each participant before the assessments commenced.

### Competing interests

The authors declare that they have no competing interests.

### Funding

This research was funded by a Future Leader in Diabetes Award from the European Foundation for the Study of Diabetes (EFSD) and the Novo Nordisk Foundation (NNF19SA058975), the Liaison Committee for education, research, and innovation in Central Norway, and the Faculty of Medicine and Health Sciences, NTNU. The sponsors have no role in study design, data collection, analysis, and publication of results.

### Authors’ contributions

TM, KÅS, and GH conceived and developed the study and analysis plan. HMSS, MAJS, KLH, and ERA collected exercise and body composition data. SS, KD, and HM performed ultrasound assessments. HMSS analysed the data and drafted the manuscript with contributions from SS. All authors reviewed the manuscript, provided feedback and approved the final manuscript.

## Acknowledgements

We would like to thank our participants for their valuable contribution to our research. We would also like to thank the Department of Obstetrics and Gynaecology at St. Olav’s University Hospital for providing facilities for exercise testing, and NeXt Move (NTNU) for providing equipment and facilities for estimation of body composition.

## Notes

### Competing Interest Statement

The authors have declared no competing interest.

### Clinical Trial

NCT04288479

### Author Declarations

The Norwegian Regional Committees for Medical and Health Research Ethics gave ethical approval for this work (ID: 62993).

