## Supplementary Table for "Acute effect of high-intensity interval training on fetal blood flow distribution"

**Supplementary Table 1. Self-reported exercise characteristics and medication use.**

| **Do you take any daily medications?** | |
| --- | --- |
| Yes | 8 (24 %) |
| No | 26 (76 %) |
| **Exercise days per week** | |
| 0 days | 4 (12%) |
| 1-2 days | 13 (46 %) |
| 3-5 days | 7 (25 %) |
| 6-7 days | 4 (14 %) |
| **Exercise duration** | |
| No exercise | 8 (27%) |
| < 30 minutes | 0 (0%) |
| 31-60 minutes | 23 (69 %) |
| > 60 min | 1 (3 %) |
| **Exercise intensity** | |
| Light | 0 (0%) |
| Moderate | 18 (75%) |
| High | 6 (25%) |

Data are presented as number of participants (%). Only participants who reported regular exercise during the past two weeks completed the follow-up questions regarding exercise days, duration and intensity.

**Supplementary table 2. Maternal and fetal heart rate responses during exercise.**

|  | n | Mean (SD) |
| --- | --- | --- |
| **Maternal heart rate (HR), beats per minute** | | |
| Warm-up | 33 | 118.3 (20.2) |
| Work-bout 1 | 32 | 150.2 (16.0) |
| Work-bout 2 | 33 | 155.0 (13.7) |
| Work-bout 3 | 33 | 158.5 (9.8) |
| Work-bout 4 | 34 | 160.1 (10.0) |
| Work-bout 5 | 34 | 160.9 (12.6) |
| Work-bout 6 | 34 | 159.3 (27.6) |
| Work-bout 7 | 34 | 164.6 (10.3) |
| Work-bout 8 | 34 | 166.6 (9.4) |
| Average HR | 34 | 159.9 (9.4) |
| Average HR, % of predicted HR maximum | 34 | 84 % (5 %) |
| Peak HR | 34 | 168.47 (8.5) |
| Peak HR, % of predicted HR maximum | 34 | 88 % (5 %) |
| Predicted HR maximum | 34 | 190.8 (2.6) |
| **Fetal heart rate, beats per minute** | | |
| Warm-up | 31 | 146.3 (9.7) |
| Work-bout 1 | 32 | 147.5 (12.0) |
| Work-bout 2 | 33 | 148.9 (13.2) |
| Work-bout 3 | 34 | 151.0 (13.3) |
| Work-bout 4 | 33 | 152.2 (11.5) |
| Work-bout 5 | 29 | 152.9 (16.4) |
| Work-bout 6 | 33 | 158.0 (16.9) |
| Work-bout 7 | 32 | 156.7 (14.3) |
| Work-bout 8 | 32 | 153.7 (15.6) |
| Average HR | 34 | 152.3 (11.9) |
| HR variability | 34 | 26.2 (13.4) |
| Peak HR | 34 | 164.9 (14.7) |
| Minimum HR | 34 | 141.2 (10.6) |
| **Rate of perceived exertion (RPE), 6-20** | | |
| Work-bout 1 | 33 | 14.3 (1.8) |
| Work-bout 2 | 34 | 14.8 (1.7) |
| Work-bout 3 | 34 | 15.4 (1.6) |
| Work-bout 4 | 32 | 15.9 (1.6) |
| Work-bout 5 | 34 | 16.1 (1.4) |
| Work-bout 6 | 34 | 16.3 (1.3) |
| Work-bout 7 | 34 | 16.5 (1.3) |
| Work-bout 8 | 34 | 16.9 (1.2) |
| Average RPE | 34 | 15.8 (1.3) |
| Peak RPE | 34 | 17.0 (1.2) |

Data are presented as mean (SD).

**Supplementary table 3. Birth and delivery outcomes.**

|  | *n* | Mean (SD) |
| --- | --- | --- |
| Difference between estimated and actual delivery date, days | 34 | -1.8 (8.9) |
| Birth weight, g | 34 | 3481 (403) |
| Head circumference, cm | 34 | 35.1 (1.6) |
| Birth length, cm | 33 | 49.8 (1.9) |
| Placenta weight, g | 33 | 586.1 (97.1) |
| **Mode of delivery, n (%)** | | |
| Vaginal delivery, n (%) | 29 (85%) | |
| Caesarean section, n (%) | 5 (15%) | |
| **Adverse events** | | |
| Preterm delivery | 1 (14%) | |
| Placenta previa | 1 (14%) | |
| Apgar score < 7 at 5 minutes | 1 (14%) | |
| Retained placenta | 1 (14%) | |
| Prolonged labor | 1 (14%) | |
| **Parity, n (%)** | | |
| Para 0 | 20 (59%) | |
| Para 1 | 12 (35%) | |
| Para 2 | 2 (6%) | |

Data are presented as mean (SD) or as n (%).
